# The relative infectiousness of asymptomatic SARS-CoV-2 infected persons compared with symptomatic individuals: A rapid scoping review

**DOI:** 10.1101/2020.07.30.20165084

**Authors:** David Mc Evoy, Conor G. McAloon, Áine B. Collins, Kevin Hunt, Francis Butler, Andrew W. Byrne, Miriam Casey, Ann Barber, John Griffin, Elizabeth Ann Lane, Patrick Wall, Simon J. More

## Abstract

**Objectives:** The aim of this study was to conduct a scoping review of estimates of the relative infectiousness of asymptomatic persons infected with SARS-CoV-2 compared with symptomatic individuals.

**Design:** Rapid scoping review of literature available until 8^th^ April 2020.

**Setting:** International studies on the infectiousness of individuals infected with SARS-CoV-2

**Participants:** Studies were selected for inclusion if they defined asymptomatics as a separate cohort distinct from pre-symptomatics and if they provided a quantitative measure of the infectiousness of asymptomatics relative to symptomatics.

**Primary outcome measures:** The relative number of secondary cases produced by an average primary case, the relative probability of transmitting infection upon contact, and the degree of viral shedding.

**Results:** Very few studies reported estimates of relative infectiousness of asymptomatic compared with symptomatic individuals. Significant differences exist in the definition of infectiousness. Viral shedding studies in general show no difference in shedding levels between symptomatic and asymptomatic individuals but are likely to be impacted by insufficient statistical power. Two contact tracing studies provided estimates of 0.7 and 1.0, but differences in approach and definition preclude comparison across the two studies. Finally, two modelling studies suggest a relative infectiousness of around 0.5 but one of these was more reflective of the infectiousness of undocumented rather than asymptomatic cases. Importantly, one contact tracing study showing a very low level of infectiousness of asymptomatic was not included in the analysis at this point due difficulties interpreting the reported findings.

**Conclusions:** The present study highlights the need for additional studies in this area as a matter of urgency. For the purpose of epidemiological modelling, we cautiously suggest that at present, asymptomatics could be considered to have a degree of infectiousness which is about 0.40-0.70 that of symptomatics. However, it must be stressed that this suggestion comes from a very low evidence base and that estimates exist that are close to zero and close to 1.

**ARTICLE SUMMARY:** *Strengths and limitations of this study:*

- Differences in the definition of infectiousness and a low number of studies estimating this parameter negate the potential to provide a pooled quantitative estimate or relative infectiousness.
- The present study highlights the need for additional studies in this area as a matter of urgency.
- Several of the studies reviewed are in pre-print stage and are not peer-reviewed.

## INTRODUCTION

The first case of a novel coronavirus (COVID-19) was first reported from Wuhan, China in December 2019. [1] The outbreak of COVID-19 was declared a Public Health Emergency of International Concern on 30 January 2020 and a pandemic was declared on 11 March 2020. [2] Since then, many countries have sought to contain the spread of the virus through a range of measures aimed at limiting transmission within the population.

At the outset of an epidemic, a key principle of control might be quarantining of individuals with clinical symptoms fitting a particular case definition. However, for many infectious diseases, a proportion of infected individuals may never present with clinical signs (i.e. asymptomatic), yet still be infectious to others. The existence of this cohort of SARS-CoV-19-infected individuals has been shown. [3]

The transmission potential of such asymptomatic individuals is likely to be different to those that have clinical signs. On the one hand, they might shed lower quantities of the infectious agent; on the other hand, their potential for contacts might be greater. Being unaware that they are infected, asymptomatic people are less likely to follow quarantine guidelines designed to restrict transmission from infected individuals.

Decision-making in the midst of a pandemic often relies on predicted outcomes from infectious disease models. Such models may aid in public health decision making by predicting the number of new cases each day as well as possible trajectories of an outbreak given different management options. Estimates from these models may be sensitive to the way in which asymptomatic individuals are considered. [4] In particular, it is important to understand the proportion of individuals who are infectious but remain asymptomatic, as well as understanding the transmission potential in that cohort, compared with symptomatic individuals, i.e. the relative infectiousness.

The purpose of this study, was to conduct a scoping review of available literature to answer the question ‘In patients infected with SARS-CoV-2 (Population), how infectious are asymptomatic individuals compared with symptomatic individuals?’ A scoping review rather than a systematic review was undertaken given the rapidly evolving and complex literature available on SARS-CoV-2 infection, and the range of ways in which ‘infectiousness’ might be defined.

## MATERIALS AND METHODS

For the purpose of this study, we defined asymptomatics as individuals who would never develop symptoms of the infection. We considered that symptomatic infection incorporated pre-symptomatic and symptomatic phases. We followed the preferred reporting items for systematic reviews and meta-analysis with extension for scoping reviews (PRISMA – ScR) guidelines.[5] A review protocol was established for this work. However, the review was not pre-registered.

### Search methodology, initial screening and categorisation

A systematic survey of the literature between 1 December 2019 and 8th April 2020 for all countries was implemented using the following search strategy. Publications on the electronic databases PubMed, Google Scholar, MedRxiv and BioRxiv were searched with the following keywords: (“Novel coronavirus” OR “SARS□CoV□2” OR “2019-nCoV” OR “COVID-19”) AND (“infectious” OR “infectiousness” OR “viral load” OR “transmission” OR “asymptomatic”). The dynamic curated PubMed database “LitCovid” [6] was also monitored, in addition to national and international government reports. No restrictions on language or publication status were imposed so long as an English abstract was available. Articles were evaluated for data relating to the aim of this review; publications were considered relevant for possible inclusion if they contained information relating to the infectiousness of asymptomatic individuals. Bibliographies within these publications were also searched for additional resources. The search was not limited to peer-reviewed sources, but also included pre-print articles available on the MedRxiv and BioRxiv databases. Initial screening was conducted on the eligibility screening by three of the co-authors (ÁC, KH, FB).

### Further screening and data extraction

Next, two authors (DME; CMA) reviewed the shortlisted studies in turn. The following data were extracted from each study: author, year, location, method of estimating relative infectiousness and data relating to the infectiousness of asymptomatics relative to symptomatics. The estimate and associated confidence intervals of this proportion were extracted. Data extraction was conducted independently by each of these authors and disparities were resolved afterwards by discussion between four of the co-authors with in-depth knowledge of the studies (DME, ÁC, CMA, SM). Authors of studies were contacted when there was some confusion over data reported. Bias was assessed qualitatively, by evaluating the methods used in each of the enrolled studies.

## RESULTS

After initial screening, 18 articles were identified as being relevant to the review question:

- Two studies [7,8] were identified as review rather than primary studies. Neither of these studies sought to quantify the relative infectiousness. Both were removed after screening their reference lists for additional studies not included in our original search.
- Three articles were identified that based their analyses on the same primary data. [9-11] Only one of these studies [10] was included, the other two studies were removed. [9,11]
- One case report [12] was removed since it did not seek to answer the question of relative infectivity, but instead presented the results of contacts of a single infected case.
- One study [13] was removed since there was not a clear definition of an ‘asymptomatic’ individuals, and, although no differences were reported in viral load according to disease severity, the profile of asymptomatic patients was not explicitly described.
- One study [14] was removed as there was difficulty in following the methods reported in that manuscript. The authors of the present study could not come to a consensus on the precise outcomes of the study.
- Three papers [4, 15, 16] were removed as they used infectiousness as input parameters in their models, rather than estimating the parameter itself.
- One study [17] was removed as the definition of asymptomatic was more closely aligned with our definition of pre-symptomatic. Furthermore, this study did not seek to estimate the infectiousness of this cohort rather evaluate the impact of different assumptions of the infectiousness of this cohort on disease transmission.
- One study [18] was removed since it combined priors to infer relative infectiousness and did not estimate the parameter from data.

Of the remaining studies (n = 6), three methods of estimating relative infectiousness (RI) were identified: two studies [19, 20] reported viral shedding loads, two studies [21, 22] reported the outputs of mathematical models, one study [13] reported the outcome of a contact tracing study and one study [23] reported both viral shedding load outcome and the results of contact tracing. Table 1 summarises the relevant values for relative infectiousness.

**Table 1.**
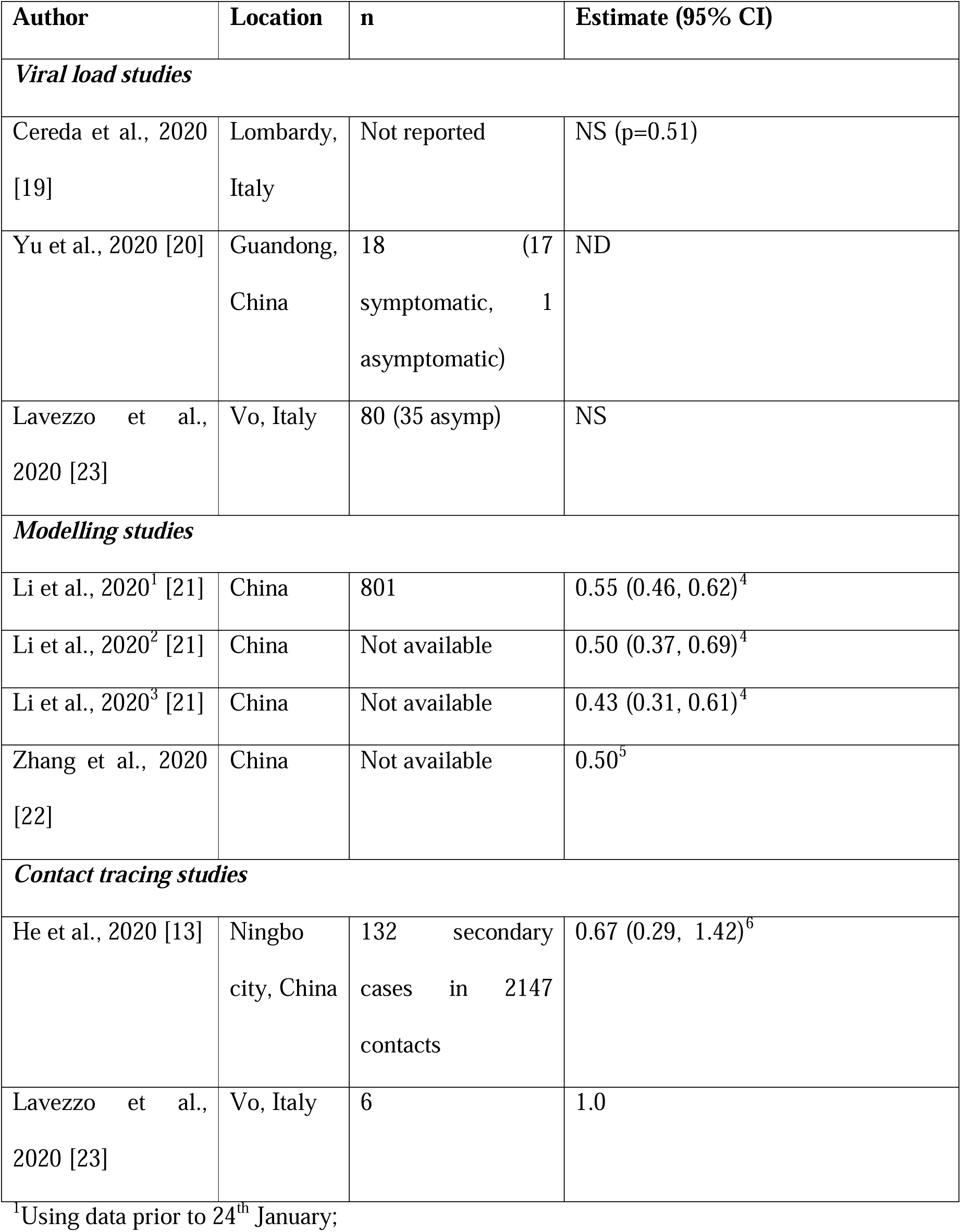
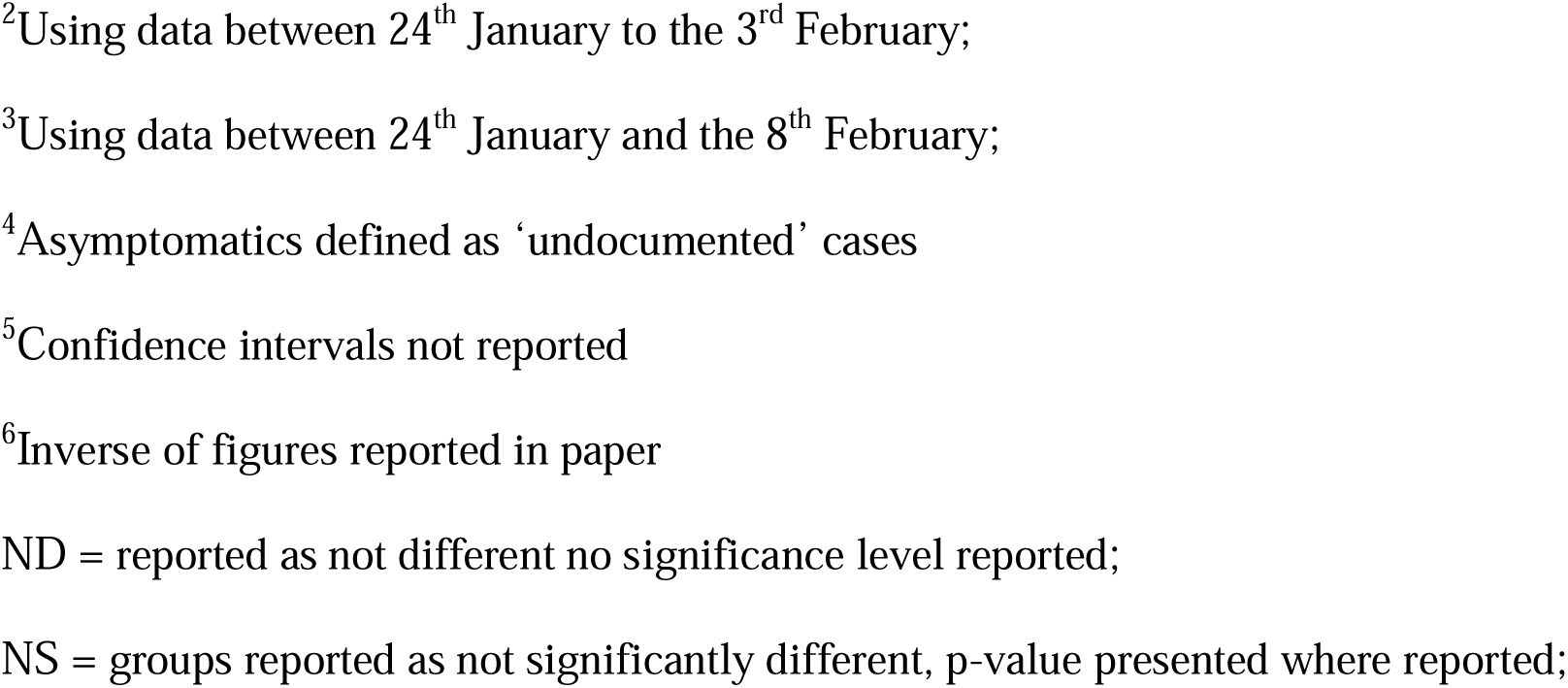
Summary of values relevant to the relative infectiousness of asymptomatic individuals.

Of the 2 contact tracing studies, 1 [13] reported that the relative risk of becoming infected after being exposed to a symptomatic individual was 1.5 (0.7 - 3.4) that of becoming infected following exposure to an asymptomatic person. The inverse of these values represents the relative risk of infection following exposure to an asymptomatic versus symptomatic individual (0.67, 95% CI: 0.29 - 1.42). The second contact tracing study [23] found that new infections in Vo’, Italy, were as likely to be attributed to asymptomatic individuals as they were symptomatic individuals. It should also be noted that this was based on a small number of samples: whilst 8 new infections occurred over the observation period in the Italian study, the source of 6 could be ascertained, with 3 attributed to symptomatics, and 3 attributed to asymptomatics.

Three viral load studies [19, 20, 23] were selected for extraction. None of these studies reported a significant difference in the viral load of asymptomatics versus symptomatics. It is important to note the number of individuals in these studies was in general small or not reported which may have affected the statistical power.

Finally, estimates were available from 2 modelling studies [21, 22]. The first of these [21] reported three estimates of the infectiousness of ‘undocumented’ infections rather than asymptomatic infections, ranging from 0.43 to 0.55. Estimates were reported across a number of different stages of the outbreak. The value reported from the most recent stage of the outbreak was also the lowest (0.43). [21] A second modelling study [22] estimated the relative infectiousness of asymptomatic at 0.5 but did not report confidence intervals.

## DISCUSSION

Determining the infectiousness of asymptomatics is important in informing public health decision making in the midst of a pandemic. Even if asymptomatic individuals are a smaller proportion of the overall cohort of infected individuals, their potential to transmit could be significant since they are unlikely to undertake the same controls (e.g. self-isolation) to limit the spread of infection to others as they are unlikely to be aware that they are infected.

Overall, at present, few studies are available to provide a robust quantitative estimate of the relative infectiousness of asymptomatic and symptomatic individuals infected with SARS-CoV-2. A significant issue encountered in this review was the degree to which the definition of infectivity differed. Inferences from contact tracing studies in particular need to be interpreted with caution. We identified two contact tracing studies [13, 23] with infectiousness values of 0.67 and 1.0. However, the two studies were based on opposing perspectives: one presented the risk of infection given exposure, whilst the other presented the risk of exposure type given infection. It is worth noting that one contact tracing study [14] was identified that reported odds of infection from an asymptomatic case that was 0.06 that of moderate symptomatic case. However, due to the reporting in that paper, the group of co-authors in the present study could not reach a consensus on exactly how the figures were determined and this study was removed from the review at that stage.

Studies reporting viral loads [refs] have generally concluded that there is no significant difference between asymptomatic and symptomatic individuals. However, these studies are generally based on a small numbers of cases and are therefore likely to be underpowered in detecting differences between these groups. Interestingly, examination of viral shedding data from two viral studies which presented raw data [19, 23] demonstrates that loads were numerically lower in asymptomatic patients. Potentially, statistically significant differences could have been detected with a greater number of observations.

Furthermore, viral shedding studies are also often cross-sectional. In such study designs, unless there is sufficient patient follow up, pre-symptomatics (that is, those individuals that do not yet have clinical signs but will go on to develop symptoms) are likely to be included in the definition of asymptomatics. However, these are two distinct cohorts from both modelling and biological perspectives. Finally, viral load is not directly relatable to infectiousness of the individual. It is likely for example that the behaviour of symptomatics and asymptomatics will differ, and therefore the opportunity to transmit infection will be different, independent of the level of viral shedding. Furthermore, many of the physical manifestations of respiratory disease, such as coughing, might also serve to disperse virus more efficiently, again independent of the degree of viral shedding as measured by PCR.

Estimates from modelling studies provide the third method of estimating relative infectiousness for this virus. Li et al. reported the infectiousness of undocumented rather than asymptomatics. [21] Three separate estimates of the infectiousness of undocumented infections were reported for the Li study. The proportion of undocumented cases (compared to documented) consecutively decreased with time points corresponding to greater restrictions. The authors argue that this was likely to have occurred since, with greater restrictions, it was more likely that symptomatic individuals would be tested and therefore become documented. Consequently, we recommend that the most recent value be used from that study, that is 0.43 (0.31, 0.61). [21]

The present study highlights the need for additional studies in this area as a matter of urgency. Taking the estimates presented together we cautiously suggest that asymptomatics could be considered to have a degree of infectiousness which is about 0.4 - 0.7 that of symptomatics. However, it must be stressed that this suggestion comes from a very low evidence base and that estimates exist that are close to zero [14] and close to 1 [23]. Previously, Ferguson et al. [15] assumed that symptomatics were 50% more infectious than asymptomatics. When converted to a ratio (i.e. 1/1.5), this corresponds to a relative infectiousness of 0.67. Tuite et al. [16] did not model asymptomatics as a distinct cohort to symptomatics. Finally, Aguilar et al. [4] used a figure for relative infectiousness of 0.55, based on the estimates of Li et al. [21]

Modelling studies requiring informative estimates of the relative infectiousness of asymptomatic individuals should seek to ensure that the precise definition of the estimate used equates to the same definition used in the model. Some definitions may be more population specific than others. In particular it is important to note whether the definition of infectiousness incorporates contact rates (which might be different for symptomatic or asymptomatic individuals), or is independent of it; or whether it incorporates the proportion of asymptomatic individuals in the population, or is independent of it.

## Conclusion

Overall, few studies were available to provide a quantitative estimate of the relative infectiousness of asymptomatics. Three approaches to estimating RI were identified that might help indicate the value for this parameter. However, there are issues with each of these approaches with respect to informing the parameter of interest. Taking the estimates from two modelling studies and one contact tracing study together, we cautiously suggest that for the purpose of modelling studies, asymptomatics could, at present, be considered to have a degree of infectiousness which is about 0.4-0.7 that of symptomatics. However, it must be stressed that this suggestion comes from a very low evidence base and that estimates exist that are close to zero and close to 1.

## Data Availability

This was a review of pre print and peer reviewed material available for researchers online

## List of abbreviations

PCR: Polymerase chain reaction
RT PCR: reverse transcription polymerase chain reaction
RI: Relative infectiousness of asymptomatic versus symptomatic infected persons with COVID-19

## Declarations

### Ethics approval and consent to participate

*not applicable*

### Consent for publication

*not applicable*

### Availability of data and materials

*not applicable*

### Patient and Public Involvement

No patient involved.

### Author contributions

All authors were involved in searching databases. DMcE, ÁC, CMA and SM drafted the first document. All authors participated in the intellectual content analysis, interpretation of the findings, and review of the final version of the document. All authors read and approved the final document.

## Acknowledgements

We wish to thank Dr Gerald Barry and Emma O’ Donohue for their feedback and input.

## REFERENCES

1. Zhou C. Evaluating new evidence in the early dynamics of the novel coronavirus COVID-19 outbreak in Wuhan, China with real time domestic traffic and potential asymptomatic transmissions. medRxiv. 2020:2020.02.15.20023440.

2. Aguilar JB, Faust JS, Westafer LM, Gutierrez JB. Investigating the Impact of Asymptomatic Carriers on COVID-19 Transmission. medRxiv. 2020:2020.03.18.20037994.

3. Hu Z, Song C, Xu C, G. Jin. Clinical characteristics of 24 asymptomatic infections with COVID-19 screened among close contacts in Nanjing, China. Sci China Life Sci. 2020;63(5):706–711. doi:10.1007-s11427-020-1661-4.

4. Sun, T, Weng, D. Estimating the effects of asymptomatic and imported patients on COVID-19 epidemic using mathematical modeling. J Med Virol. 2020; 1– 9. https:--doi.org-10.1002-jmv.25939.

5. Tricco AC, Lillie E, Zarin W, O’Brien KK, et al. PRISMA extension for scoping reviews (PRISMA-ScR): checklist and explanation. Annals of internal medicine. 2018 Oct 2;169(7):467–73.

6. LitCovid. LitCovid 2020 [Available from: https://www.ncbi.nlm.nih.gov-research-coronavirus-.

7. Aguirre-Duarte N, 2020. Can people with asymptomatic or pre-symptomatic COVID-19 infect others: a systematic review of primary data. medRxiv.

8. Gao Z, Xu Y, Sun C, Wang X, Guo Y, Qiu S, Ma K. A Systematic Review of Asymptomatic Infections with COVID-19. Journal of Microbiology, Immunology and Infection. 2020 May 15.

9. Chen Y, Wang A, Yi B, Ding K, Wang H, Wang J, et al. The epidemiological characteristics of infection in close contacts of COVID-19 in Ningbo city. Chin J Epidemiol 2020;41.

10. He D, Zhao S, Lin Q, Zhuang Z, Cao P, Wang MH, Yang L. The relative transmissibility of asymptomatic cases among close contacts. International Journal of Infectious Diseases. 2020 Apr 18.

11. Yin G, Jin H. Comparison of transmissibility of coronavirus between symptomatic and asymptomatic patients: Reanalysis of the Ningbo Covid-19 data. medRxiv. 2020 Jan 1.

12. Gao M, Yang L, Chen X, Deng Y, et al., 2020 A study on infectivity of asymptomatic SARS-CoV-2 carriers. Respiratory Medicine. 2020 May 13:106026.

13. He X, Lau EH, Wu P, Deng X, et al., 2020. Temporal dynamics in viral shedding and transmissibility of COVID-19. Nature medicine. 2020 Apr 15:1-4.

14. Luo L, Liu D, Liao X-l, Wu X-b, Jing Q-l, Zheng J-z, et al. Modes of contact and risk of transmission in COVID-19 among close contacts. medRxiv. 2020:2020.03.24.20042606.

15. Ferguson N, Laydon D, Nedjati-Gilani NI G, Ainslie K, Baguelin M, et al. Impact of non-pharmaceutical interventions (NPIs) to reduce COVID-19 mortality and healthcare demand. Imperial College, London. 2020.

16. Tuite A, Fisman DN, Greer AL. Mathematical modeling of COVID-19 transmission and mitigation strategies in the population of Ontario, Canada. medRxiv. 2020:2020.03.24.20042705.

17. Mandal S, Bhatnagar T, Arinaminpathy N, Agarwal A, Chowdhury A, Murhekar M, et al. Prudent public health intervention strategies to control the coronavirus disease 2019 transmission in India: A mathematical model-based approach. The Indian journal of medical research. 2020.

18. Ferretti L, Wymant C, Kendall M, Zhao L, Nurtay A, Abeler-Dorner L, et al. Quantifying SARS-CoV-2 transmission suggests epidemic control with digital contact tracing. medRxiv. 2020:2020.03.08.20032946.

19. Cereda DT, M; Rovida, F; Demicheli, V; Ajelli, M, et al. The early phase of the COVID-19 outbreak in Lombardy, Italy. 2020. arXiv preprint arXiv:2003.09320.

20. Yu J, Yen H-L, Huang H, Wu J, Ruan F, Liang L, et al. SARS-CoV-2 Viral Load in Upper Respiratory Specimens of Infected Patients. The New England Journal of Medicine. 2020;382(12):1177–9.

21. Li R, Pei S, Chen B, Song Y, Zhang T, Yang W, et al. Substantial undocumented infection facilitates the rapid dissemination of novel coronavirus (SARS-CoV2). Science (New York, NY). 2020:eabb3221.

22. Zhang Y, You C, Cai Z, Sun J, Hu W, Zhou X-H. Prediction of the COVID-19 outbreak based on a realistic stochastic model. medRxiv. 2020:2020.03.10.20033803.

23. Lavezzo E, Franchin E, Ciavarella C, Cuomo-Dannenburg G, et al. Suppression of COVID-19 outbreak in the municipality of Vo, Italy. medRxiv. 2020.04.17.20053157; doi: https:--doi.org-10.1101-2020.04.17.20053157

